# Impact of Childhood Household Support on Depression and Self-Reported Mental and Physical Health

**DOI:** 10.1101/2025.07.03.25330796

**Authors:** Oluwasegun Akinyemi, Mojisola Fasokun, Fadeke Ogunyankin, Raolat Adenike Salami, Ifunanya Stella Osondu, Akachukwu N. Eze, Kakra Hughes, Miriam Michael, Temitope Ogundare

**Affiliations:** The Clive O Callender Outcomes Research Center, Howard University College of Medicine, Washington DC, USA; Department of Epidemiology, University of Alabama at Birmingham, AL, USA; Department of Research Data Science and Analytics, Cook Children’s Health Care System: Cook Children’s Medical Center, Fort Worth., Texas, USA; Department of Medicine and Health Sciences, Afe Babalola University, Ado-Ekiti, Ekiti State, Nigeria; Department of Public Health, Western Illinois University, Macomb, IL, USA; Department of Surgery, Howard University College of Medicine, Washington DC, USA; Department of Internal Medicine, Howard University College of Medicine Washington, DC, USA; Department of Psychiatry, Boston University School of Medicine Boston, Massachusetts, USA

**Keywords:** perceived household support, childhood experiences, depression, mental health, physical health, BRFSS

## Abstract

**Background:** Perceived household support during childhood may have long-term effects on mental and physical health across the life course. However, the specific associations between early supportive environments and adult health outcomes remain underexplored.

**Methods:** We conducted a cross-sectional analysis using data from the Behavioral Risk Factor Surveillance System (BRFSS) collected between 2016 and 2023. The study included 31,233 U.S. adults aged 18 years and older who provided complete responses regarding perceived childhood household support, depression diagnosis, and the number of poor mental and physical health days. The primary exposure was self-reported childhood support, categorized as: “Never,” “A Little of the Time,” “Some of the Time,” “Most of the Time,” or “All of the Time.” Outcomes included lifetime diagnosis of depression, average monthly poor mental health days, and poor physical health days. Analyses were adjusted using inverse probability weighting and controlled for sociodemographic factors, survey weights, and state, year, and month fixed effects.

**Results:** Among respondents (mean age 52.2 years; 63.4% female; 76.0% White), individuals who reported “Never” being supported during childhood were 19.4 percentage points more likely to report a depression diagnosis (95% CI: 11.6–27.2), experienced 5.33 more poor mental health days (95% CI: 3.64–7.03), and 2.77 more poor physical health days per month (95% CI: 1.23– 4.32), compared to those who reported being “Always” supported. A clear dose-response relationship was observed across all categories of household support.

**Conclusions:** Lower levels of perceived childhood household support are significantly associated with increased risk of adult depression and greater burden of poor mental and physical health. Interventions targeting early supportive environments may improve population health outcomes across the life span.

**Key Points:** *Question:* Is perceived childhood household support associated with depression and self-reported mental and physical health outcomes in adulthood?

*Findings:* In this cross-sectional study of 31,233 U.S. adults, individuals reporting they were never supported during childhood had significantly higher depression risk and reported poorer mental and physical health days compared to those always supported, with results showing a consistent gradient across varying support levels.

*Meaning:* These findings suggest policies promoting consistent childhood household support may enhance lifelong mental and physical health outcomes.

## INTRODUCTION

Childhood household support plays a pivotal role in shaping long-term health and well-being[1] and is often defined by the presence of an adult who ensures that basic needs are met and provides a stable environment for growth.[2–4]

A supportive childhood environment has been shown to be critical for fostering healthy development, while a lack of perceived household support can have profound and enduring consequences on emotional, physical, psychological and emotional well-being ranging from mental health outcomes such as depression to physical health outcomes such as heart disease.[5–7] Conversely, adverse childhood experiences, such as parental death, parental separation or divorce and parental incarceration, can create a non-supportive environment which can be associated with significant psychological and social challenges such as depression, anxiety, social isolation, impaired relationships, and reduced quality of life.[8–10]

Extensive research has established that adverse childhood experiences are linked to a higher likelihood of mental health disorders, including depression, anxiety, and reduced physical well-being in adulthood.[11, 12] The absence of consistent household support during childhood may lead to heightened vulnerability to these outcomes by disrupting emotional regulation and coping mechanisms.[13] Furthermore, childhood instability can have life-long implications for marital stability, as early experiences often shape interpersonal relationships and the capacity to build and sustain partnerships in adulthood.[14, 15]

Depression, as a leading cause of disability globally[16], is a significant public health concern with poor mental health contributing to considerable economic, emotional, and social burdens.[17, 18] Previous studies have highlighted the impact of adverse childhood experiences on health outcomes such as depression, chronic pain and other chronic physical conditions[19], with low socioeconomic status and adverse childhood experiences appearing to both lead to adverse health; as such, impoverished adults with a history of childhood adversity, as compared to their wealthier peers, may be at differentially increased risk for these poor health outcomes.[20]

However, the unique contribution of childhood household support has not been thoroughly investigated.[21, 22] This study seeks to address this gap by examining the life-long impact of perceived childhood household support on depression and self-reported mental and physical health.

## METHODOLOGY

### Study Design and Data Source

This study utilized data from the Behavioral Risk Factor Surveillance System (BRFSS)[23, 24] collected between January 2016 and December 2023.[25] The BRFSS is a nationally representative, cross-sectional survey conducted annually by the Centers for Disease Control and Prevention (CDC).[21] It employs a complex multistage sampling design incorporating stratification, clustering, and unequal probabilities of selection. The survey is administered via landline and cellular telephone interviews across all 50 U.S. states, the District of Columbia, and U.S. territories. Survey weights are applied to adjust for non-response, sampling design, and demographic post-stratification, ensuring generalizability to the U.S. non-institutionalized adult population.[23]

### Study Population

The target population for this study included adults aged 18 years and older residing in the United States who participated in the BRFSS survey between 2016 and 2023. Respondents were included in the analytic sample if they had complete responses to the question the survey question: “For how much of your childhood was there an adult in your household who tried hard to make sure your basic needs were met? to measure the key exposure variable (the presence of childhood household support), all three outcome variables (depression diagnosis, mental health days, and physical health days), and all relevant covariates. To preserve analytic integrity and ensure comparability, individuals were excluded if they responded, “Don’t know/Not sure,” “Refused,” or provided missing responses to any variable of interest. After applying these inclusion and exclusion criteria, the final analytic sample consisted of 31,233 respondents.

### Primary Explanatory Variables

The primary explanatory variables were measures of perceived childhood household support. This was assessed using the survey question: “For how much of your childhood was there an adult in your household who tried hard to make sure your basic needs were met?” Responses included: never, a little of the time, some of the time, most of the time, or all of the time and the treated group consisted of those that responded with - never, a little of the time, some of the time, most of the time and compared with the control group who responded with - all of the time.

This variable was used to categorize the level of perceived childhood household support. Responses of “Don’t know/not sure,” “Refused,” or missing values were excluded from the analysis.

### Outcome Variables

The study examined three primary outcomes: depression, self-reported mental health and self-reported physical health.

Depression: Measured by the question, “(Ever told) (you had) a depressive disorder (including depression, major depression, dysthymia, or minor depression).” Responses were categorized into a binary variable (1 = diagnosis of depression, 0 = no diagnosis of depression).

Mental Health: Derived from the question, “Now thinking about your mental health, which includes stress, depression, and problems with emotions, for how many days during the past 30 days was your mental health not good?” Responses ranged from 0 to 30 days.

Physical Health: Based on the question, “Now thinking about your physical health, which includes physical illness and injury, for how many days during the past 30 days was your physical health not good?” Responses ranged from 0 to 30 days.

Responses of “Don’t know/not sure,” “Refused,” or missing values were excluded from the analysis for all outcome variables.

### Covariates and Coding

Covariates were selected based on known associations with childhood household support and health outcomes. Age was included as a continuous variable. Gender was coded as male or female. Race/ethnicity was grouped into four categories: non-Hispanic White, non-Hispanic Black, Hispanic, and Other. Education level was categorized as high school or less, college degree or advanced degree. Marital status was coded as married, divorced, separated, never married, widowed or member of an unmarried couple. Employment status included employed or unemployed. Insurance type included private, Medicare, Medicaid, self-pay, and other. Metropolitan status distinguished between metro and non-metro areas, and urbanicity was coded as urban or rural. Language spoken at home was categorized as English or Spanish. State and year of survey were included as fixed effects. All categorical covariates were dummy coded for inclusion in modeling.

### Handling of Missing Data

We addressed missing data using complete case analysis, restricting our analytical sample to participants with complete information on all study variables. This approach was deemed appropriate after confirming that the data were missing at random (MAR), as missingness was systematically related to observed covariates included in our analytical models. Therefore, the complete case analysis ensured valid inference without introducing significant bias due to missingness patterns.

### Statistical Analysis

We employed inverse probability weighting to estimate the average treatment effects of perceived childhood household support on depression, poor mental health days, and poor physical health days. This approach approximates a randomized trial by weighting individuals based on the inverse of their probability of exposure, thereby balancing observed covariates across childhood household support categories and reducing confounding bias in the estimation of causal effects.

### Step 1: Propensity Score Estimation

Propensity scores were estimated using a multinomial logistic regression with childhood household support categories as the outcome and all covariates as predictors:

*mlogit childhood household support age i.sex i.race i.education i.marital_status i.employment i.insurance i.metro i.urban, baseoutcome(Never) predict ps1 ps2 ps3 ps4 ps5, pr*

### Step 2: Compute Inverse Probability Weights

Inverse probability weights were calculated based on the predicted probability of each respondent’s observed childhood household support category:

*gen ipw =* .

*replace ipw = 1/ps1 if childhood household support == “Never“*

*replace ipw = 1/ps2 if childhood household support == “A Little of the Time“*

*replace ipw = 1/ps3 if childhood household support == “Some of the Time“*

*replace ipw = 1/ps4 if childhood household support == “Most of the Time“*

*replace ipw = 1/ps5 if childhood household support == “All of the Time“*

To limit the influence of extreme weights, values were trimmed at the 1st and 99th percentiles: *summarize ipw, detail*

*scalar p1 = r(p1)*

*scalar p99 = r(p99)*

*replace ipw = p1 if ipw < p1*

*replace ipw = p99 if ipw > p99*

*Step 3: Incorporate Survey Weights and Inverse Probability Weights*

To ensure national representativeness, the BRFSS sampling weight (_llcpwt) was multiplied by the IPW to generate the final analysis weight (pweight):

*gen pweight = _llcpwt * ipw*

Models were estimated using survey-weighted regression with robust standard errors clustered by primary sampling unit:

*svyset _psu [pw = pweight], strata(_ststr) singleunit(centered)*

*svy: regress mental_health_days i.childhood household support*

*svy: regress physical_health_days i.childhood household support*

*svy: logit depression i.childhood household support*

### Software and Reproducibility

All statistical analyses were performed using Stata/SE version 18.0 (StataCorp LLC, College Station, TX). Propensity scores were estimated using multinomial logistic regression, and inverse probability weights (IPW) were manually computed to simulate treatment assignment across childhood household support categories. These weights were combined with BRFSS-provided sampling weights to generate final analysis weights that accounted for both treatment selection bias and complex survey design.

Survey-adjusted regression models were fitted using the svy suite of commands with robust standard errors clustered by primary sampling unit. Post-estimation marginal effects and predicted probabilities were calculated using the margins command to quantify outcome differences across childhood household support categories.

### Ethics and Data Access

The BRFSS dataset is publicly available and de-identified, ensuring adherence to ethical standards. As the study utilized publicly available data, additional Institutional Review Board (IRB) approval was not required.

## RESULTS

Table 1 summarizes demographic and health characteristics of 31,233 U.S. adults by perceived childhood household support. Significant differences were noted across all variables. Mean age was 52.2 ± 17.7 years, with the youngest individuals among those supported “a Little of the Time” (47.1 ± 16.3 years). Most participants were women (63.4%), disproportionately represented in the “Never Supported” group (66.2%) compared to the “Always Supported” group (61.8%). White respondents represented most of the sample (76.0%), while Hispanic individuals were disproportionately found among those reporting less support (10.1% “Never Supported,” 10.6% “a Little of the Time”). Married individuals were more common among those “Always Supported” (46.6%) than “Never Supported” (33.9%). Employment was lowest among those “Never Supported” (33.5%) versus “Always Supported” (44.6%). Higher education levels correlated positively with perceived support; 69.1% of those “Always Supported” had college degrees or higher compared to 52.9% of the “Never Supported” group. Spanish speakers were more prevalent in lower-support categories (4.4% “Never Supported” vs. 1.4% “Always Supported”). Metropolitan residence was similarly distributed, though slightly less common in the “Never Supported” group (69.2%).

**Table 1:** Demographic and Health Characteristics by Levels of Childhood Household Support.

| Variable | Total Population | All of the time | Never | A little of the time | Some of the time | Most of the time | chi2 | p-value |
| --- | --- | --- | --- | --- | --- | --- | --- | --- |
|  | (N= 31,233) | (n=18,400) | (n=1,556) | (n=1,448) | (n=2,934) | (n=6,895) |  |  |
| **Age (Yr.)** | $52.2 \pm 17.7$ | $54.1 \pm 17.6$ | $52.2 \pm 17.0$ | $47.1 \pm 16.3$ | $48.7 \pm 17.5$ | $49.9 \pm 17.8$ | 22.56 | < 0.001 |
| **Sex** |  |  |  |  |  |  | 107.59 | <0.001 |
| Female | 19,809 (63.4%) | 11,374 (61.8%) | 1,030 (66.2%) | 1,027 (70.9%) | 2,038 (69.5%) | 4,340 (62.9%) |  |  |
| Male | 11,424 (36.6%) | 7,026 (38.2%) | 536 (33.8%) | 421 (29.1%) | 896 (30.5%) | 2,555 (37.1%) |  |  |
| **Race & Ethnicity** |  |  |  |  |  |  | 216.<br>19 | <0.00<br>1 |
| White | 16,315<br>(76.0%) | 9,793<br>(76.8%) | 741<br>(69.8%) | 721 (73.7%) | 1,531<br>(76.2%) | 3,529<br>(75.6%) |  |  |
| Black | 2,346<br>(10.9%) | 1,564<br>(12.3%) | 114<br>(10.7%) | 67 (6.8%) | 151 (7.5%) | 450 (9.6%) |  |  |
| Hispanic | 1,355 (6.3%) | 647 (5.1%) | 107<br>(10.1%) | 104 (10.6%) | 157 (7.8%) | 340 (7.3%) |  |  |
| Other | 1,445 (6.7%) | 740 (5.8%) | 99 (9.3%) | 87 (8.9%) | 171 (8.5%) | 348 (7.5%) |  |  |
| **Insurance** |  |  |  |  |  |  | 437.<br>18 | <0.00<br>1 |
| Private | 7,946<br>(42.4%) | 4,952<br>(44.4%) | 229<br>(25.9%) | 291 (34.2%) | 644 (37.7%) | 1,830<br>(44.0%) |  |  |
| Medicare | 5,825<br>(31.1%) | 3,702<br>(33.2%) | 282<br>(31.9%) | 215 (25.2%) | 464 (27.2%) | 1,162<br>(28.0%) |  |  |
| Medicaid | 1,957<br>(10.4%) | 909 (8.2%) | 171<br>(19.3%) | 147 (17.3%) | 250 (14.6%) | 480 (11.6%) |  |  |
| Self-Pay | 967 (5.2%) | 468 (4.2%) | 84 (9.5%) | 74 (8.7%) | 132 (7.7%) | 209 (5.0%) |  |  |
| Other | 2,059<br>(11.0%) | 1,123<br>(10.1%) | 119<br>(13.5%) | 125 (14.7%) | 218 (12.8%) | 474 (11.4%) |  |  |
| **Marital Status** |  |  |  |  |  |  | 478.<br>81 | <0.00<br>1 |
| Married | 13,515<br>(43.6%) | 8,517<br>(46.6%) | 522<br>(33.9%) | 531 (37.0%) | 1,090<br>(37.4%) | 2,855<br>(41.7%) |  |  |
| Divorced | 6,635<br>(21.4%) | 3,594<br>(19.7%) | 303<br>(19.7%) | 365 (25.4%) | 688 (23.6%) | 1,685<br>(24.6%) |  |  |
| Widowed | 5,114<br>(16.5%) | 2,785<br>(15.2%) | 367<br>(23.8%) | 279 (19.4%) | 563 (19.3%) | 1,120<br>(16.3%) |  |  |
| Separated | 3,302<br>(10.6%) | 2,149<br>(11.8%) | 187<br>(12.1%) | 92 (6.4%) | 268 (9.2%) | 606 (8.8%) |  |  |
| Never Married | 853 (2.8%) | 435 (2.4%) | 76 (4.9%) | 64 (4.5%) | 104 (3.6%) | 174 (2.5%) |  |  |
| Member of an Unmarried Couple | 1,614 (5.2%) | 807 (4.4%) | 87 (5.6%) | 105 (7.3%) | 200 (6.9%) | 415 (6.1%) |  |  |
| **Employed (ref. Unemployed)** | 14,066<br>(45.3%) | 8,153<br>(44.6%) | 518<br>(33.5%) | 680 (47.2%) | 1,321<br>(45.2%) | 3,394<br>(49.5%) | 142.<br>21 | <0.00<br>1 |
| **Education Level** |  |  |  |  |  |  | 376.<br>21 | <0.00<br>1 |
| High School or Less | 10,227<br>(32.8%) | 5,675<br>(30.9%) | 731<br>(47.2%) | 556 (38.6%) | 1,062<br>(36.3%) | 2,203<br>(32.0%) |  |  |
| College | 9,604<br>(30.8%) | 5,456<br>(29.7%) | 452<br>(29.2%) | 518 (35.9%) | 1,000<br>(34.2%) | 2,178<br>(31.7%) |  |  |
| Advanced | 11,326<br>(36.4%) | 7,229<br>(39.4%) | 367<br>(23.7%) | 368 (25.5%) | 863 (29.5%) | 2,499<br>(36.3%) |  |  |
| **English Speaking (ref. Spanish)** | 30,762<br>(98.5%) | 18,149<br>(98.6%) | 1,487<br>(95.6%) | 1,408 (97.2%) | 2,890<br>(98.5%) | 6,828<br>(99.0%) | 120.<br>98 | <0.00<br>1 |
| **Metro (ref. non-metro)** | 22,187<br>(71.0%) | 12,921<br>(70.2%) | 1,077<br>(69.2%) | 1,031 (71.2%) | 2,127<br>(72.5%) | 5,031<br>(73.0%) | 23.9<br>6 | <0.00<br>1 |

Table 2 illustrates the average effects of childhood household support on the likelihood of receiving a depression diagnosis in adulthood. Compared to individuals who were supported “All of the time” (reference mean = 0.389; SE = 0.01; 95% CI = 0.369–0.409), participants who reported being “Never Supported” had a significantly higher likelihood of depression (Coefficient = 0.194; SE = 0.040; t = 4.880; p < 0.001; 95% CI = 0.116–0.272). Those who experienced support only “a Little of the Time” demonstrated the greatest increased risk of depression (Coefficient = 0.233; SE = 0.040; t = 5.880; p < 0.001; 95% CI = 0.156–0.311). Similarly, individuals supported “Some of the Time” also showed significantly elevated odds of depression (Coefficient = 0.193; SE = 0.028; t = 6.970; p < 0.001; 95% CI = 0.138–0.247). Participants who received support “Most of the Time,” although having lower risks than other groups with less frequent support, still exhibited significantly higher odds of depression compared to the consistently supported reference group (Coefficient = 0.087; SE = 0.020; t = 4.270; p < 0.001; 95% CI = 0.047–0.127).

**Table 2:** Average Effects of Childhood Household Support and Depression Diagnosis.

|  | Contrast | Std. Err. | t | 95% CI |  |  |
| --- | --- | --- | --- | --- | --- | --- |
| **All of the time** | 0.389 | 0.01 | 3.806 | 0.369 | 0.409 | <0.001 |
| Never vs. All of the time | +0.194 | 0.040 | 4.880 | 0.116 | 0.272 | <0.001 |
| A little of the time vs. All of the time | +0.233 | 0.040 | 5.880 | 0.156 | 0.311 | <0.001 |
| Some of the time vs. All of the time | +0.193 | 0.028 | 6.970 | 0.138 | 0.247 | <0.001 |
| Most of the time vs. All of the time | +0.087 | 0.020 | 4.270 | 0.047 | 0.127 | <0.001 |

*This table presents the Average Treatment Effects (ATE) of childhood household support on the likelihood of depression diagnosis in adulthood. The main explanatory variable categorizes childhood household support as Never, A little of the time, Some of the time, Most of the time, or All of the time (control/ reference group).*
**Coefficients (Coef.) represent the change in depression risk relative to the reference group,** with statistical significance assessed using z-values, p-values, and 95% confidence intervals (CI). Higher coefficients indicate an increased likelihood of depression with lower levels of childhood household support.

Table 3 illustrates the association between childhood household support and the average number of poor mental health days reported per month in adulthood. Compared to individuals who were supported “All of the time” (reference mean = 11.650 days; SE = 0.213, 95% CI = 11.233– 12.068), participants who were “Never Supported” reported significantly more poor mental health days (Coefficient = 5.333; SE = 0.865; t = 6.170; p < 0.001; 95% CI = 3.638–7.029). Similarly, individuals supported “a Little of the Time” also exhibited a substantial and statistically significant increase in poor mental health days (Coefficient = 5.354; SE = 0.884; t = 6.060; p < 0.001; 95% CI = 3.621–7.087). Participants who reported receiving support “Some of the Time” experienced fewer additional poor mental health days compared to the previously mentioned groups, yet the association remained significant (Coefficient = 3.999; SE = 0.531; t = 7.540; p < 0.001; 95% CI = 2.959–5.039). Even those who received support “Most of the Time” showed a statistically significant, though smaller increase in poor mental health days relative to those always supported (Coefficient = 1.084; SE = 0.429; t = 2.530; p < 0.001; 95% CI = 0.244–1.925).

**Table 3:** Average Effects of Childhood Household Support on Self-Reported Poor Mental Health Days.

|  | Contrast | Std. Err. | t P>t | 95% CI |  |  |
| --- | --- | --- | --- | --- | --- | --- |
| All of the time | 11.650 | 0.213 | 5.469 | 11.233 | 12.068 | <0.001 |
| Never vs. All of the time | +5.333 | 0.865 | 6.170 | 3.638 | 7.029 | <0.001 |
| A little of the time vs. All of the time | +5.354 | 0.884 | 6.060 | 3.621 | 7.087 | <0.001 |
| Some of the time vs. All of the time | +3.999 | 0.531 | 7.540 | 2.959 | 5.039 | <0.001 |
| Most of the time vs. All of the time | +1.084 | 0.429 | 2.530 | 0.244 | 1.925 | <0.001 |

*This table presents the Average Treatment Effects (ATE) of childhood household support on the number of self-reported poor mental health days per month in adulthood. The explanatory variable categorizes childhood household support as Never, A little of the time, Some of the time, Most of the time, or All of the time (control/reference group). Coefficients (Coef.) represent the estimated increase in poor mental health days relative to the reference group. Statistical significance is assessed using z-values, p-values, and 95% confidence intervals (CI). Higher coefficients indicate more poor mental health days associated with lower levels of childhood household support.*

Table 4 demonstrates the association between childhood household support and the average number of self-reported poor physical health days per month in adulthood. Compared to those who were “Always Supported” (reference mean = 10.606 days; SE = 0.214, 95% CI = 10.188– 11.025), individuals who were “Never Supported” reported significantly more poor physical health days (Coefficient = 2.772; SE = 0.789; t = 3.510; p < 0.001; 95% CI = 1.226–4.319). Participants who received support only “a Little of the Time” also showed a statistically significant increase, albeit smaller (Coefficient = 1.619; SE = 0.806; t = 2.010; p = 0.045; 95% CI = 0.038–3.199). Similarly, individuals supported “Some of the Time” experienced significantly more poor physical health days compared to the consistently supported group (Coefficient = 1.155; SE = 0.547; t = 2.110; p = 0.035; 95% CI = 0.082–2.228). In contrast, those who received support “Most of the Time” did not show a significant difference in poor physical health days relative to those who were always supported (Coefficient = 0.010; SE = 0.407; t = 0.020; p = 0.981; 95% CI = -0.788–0.808).

**Table 4:**
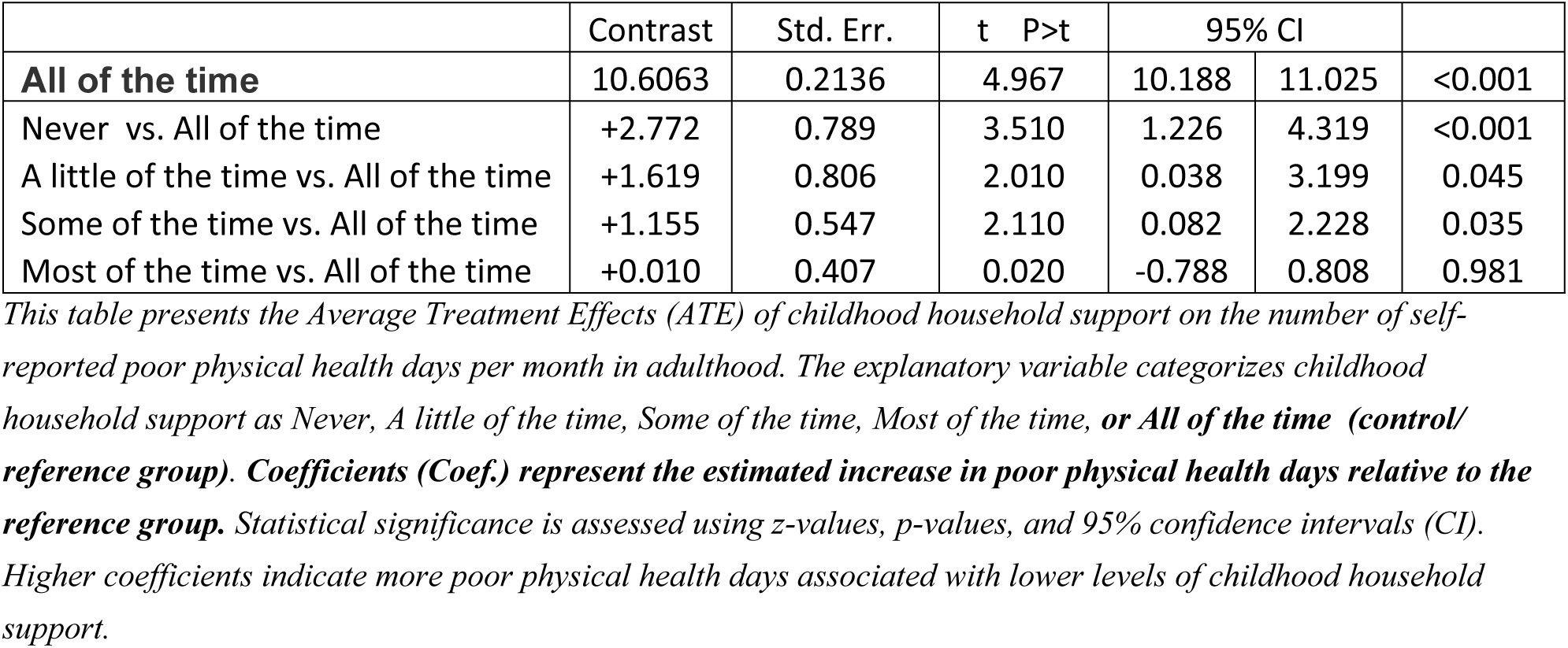
Average Effects of Childhood Household Support on Self-Reported Poor Physical Health Days.

## DISCUSSION

This study aimed to examine the relationship between perceived childhood social support and depression, poor mental health and poor physical health days in adulthood highlighting the significant role of childhood household support in shaping long-term health and relationship outcomes. The findings demonstrate that individuals with adequate childhood social support reported lower rates of depression diagnosis and fewer days of poor mental and physical health compared to those who reported poor perceived childhood social support. This was demonstrated through a linear, dose-response relationship, where the extent of perceived childhood household support corresponded to improved mental and physical well-being in adulthood.

These results aligned with the well-documented impact of adverse childhood experiences (ACEs) on adult mental health.[11, 12, 26] A nurturing childhood environment has been shown to provide emotional security, stability, and coping mechanisms that serve as protective buffers against stress and mental health disorders.[27–29] Conversely, ACEs have been associated with increased rates of mental and physical health disorders such as depression, substance use disorder, hypertension, diabetes, and obesity[29–34] as well as poor healthy behavior and difficulties in maintaining long-term relationships.[32, 33]

Increasing levels of reported perceived childhood social support was associated with progressively better outcomes, less depression, and less poor mental and physical health days. These findings support the notion that cumulative exposure to a stable and nurturing environment contributes to resilience.[35–37] According to attachment theory, secure attachment in childhood forms the foundation for emotional regulation, interpersonal relationships, and overall well-being in adulthood.[38, 39] Consistent and responsive parents or parental figures provide the developing child with a healthy sense of self, helps with regulation of the HPA axis in response to stress, and leads to adaptive neuroplastic changes which contribute to resilience and better ability to cope with stress.[36]

### Policy and Public Health Implications

The findings from this study have substantial implications for public health and policy. First, policies aimed at strengthening family structures, improving access to economic resources for single-parent households, and expanding mental health services for at-risk parents can help provide parents with resources to provide a stable and supportive environment for children. Second, investing in childhood support programs may serve as a critical preventive strategy against long-term mental and physical health burdens, especially during the early stages of childhood which is an important critical period for brain growth and maturation. Third, school-based interventions that promote emotional resilience, relationship skills, and mental health awareness may further enhance protective factors among children from less stable households. In addition, screening and identification of at-risk children and adolescents may help to initiate interventions to prevent the development of mental health conditions and to mitigate the negative effects of childhood instability. Fourth, pediatricians and clinical providers should incorporate universal screening for childhood adversity during routine visits. This will enable early detection and initiation of early intervention and support services for those at higher risk of long-term health complications.

### Limitations and Future Directions

Despite its strengths, this study has several limitations. First, the reliance on self-reported survey data introduces the potential for recall bias and subjective misclassification. Individuals may perceive and report their childhood experiences differently based on current mental states or social desirability biases. Second, despite using established causal inference methodologies, the cross-sectional study design inherently limits definitive causal conclusions. Longitudinal studies, tracking childhood experiences and subsequent adult health outcomes, would further strengthen causal evidence. Additionally, while adjustments for potential confounders were made, residual confounding due to unmeasured factors such as genetic predispositions, early-life socioeconomic conditions, or parental mental health remains possible. Future research should explore these variables in greater depth to refine the understanding of the pathways linking childhood household support to adult outcomes.

### Conclusion

This study provides robust evidence that perceived childhood household support plays a crucial role in determining adult mental and physical health outcomes, with absent childhood support associated with increased risk of depression and poorer mental and physical health. In addition, there is a dose-response relationship between the degree of perceived childhood social support and health outcomes with increasing levels of childhood social support linked to better mental and physical health and less likelihood of depression.

## Data Availability

All relevant data are within the manuscript and its Supporting Information files.

